# The Benefit of the Doubt Phenomenon in Emergency Triage Assignment Disparities

**DOI:** 10.64898/2026.02.12.26346184

**Authors:** Blanca Romero Milà, Helena Coggan, Andrew M. Fine, Yuval Barak-Corren, Ben Y. Reis, Jaya Aysola, Pradip P. Chaudhari, William G. La Cava

**Affiliations:** Computational Health Informatics Program, Boston Children’s Hospital, Boston, MA, USA; Harvard Medical School, Boston, MA, USA; Department of Biomedical Engineering, University of California, Irvine, Irvine, CA, USA; Division of Emergency Medicine, Boston Children’s Hospital, Boston, MA, USA; Division of Emergency and Transport Medicine, Children’s Hospital Los Angeles and Department of Pediatrics, Keck School of Medicine of the University of Southern California, Los Angeles, CA, USA; Department of Pediatric Cardiology, Schneider Children’s Medical Center, Affiliated to Tel Aviv University Faculty of Medical and Health Sciences, Petach Tikvah, Israel; Leonard Davis Institute of Health Economics, University of Pennsylvania, Philadelphia, PA, USA; Department of Medicine, Perelman School of Medicine, University of Pennsylvania, Philadelphia, PA, USA; Penn Medicine Center for Health Equity Advancement, Philadelphia, PA, USA

**Keywords:** emergency, triage, fairness, demographics

## Abstract

Emergency department (ED) triage decisions critically impact patient care and are standardized, yet ethnoracial disparities in triage assignment are well documented. We analyzed ethnoracial differences in triage assignments across four U.S. EDs (two adult, two pediatric), comprising 1.4 million encounters from 2011–2025. To better characterize these disparities, we developed an automated triage algorithm that replicates the Emergency Severity Index (ESI) criteria, the standard triage protocol used at each site. The algorithm identifies high-acuity symptoms and danger-zone vital signs that inform triage decisions at the level-2 (emergent) versus level-3 (urgent) boundary. We compared nurse triage assignments across ethnoracial groups, stratified by algorithmic ESI scores, using causal inference methods to adjust for clinical presentation and hospital context. Significant ethnoracial disparities in triage assignment were observed across all sites. Disparities were concentrated among patients algorithmically classified as lower risk but assigned higher acuity by nurses. This pattern is consistent with a “benefit-of-the-doubt” disparity, in which relatively stable, non-Hispanic White patients are more often assigned higher priority than Hispanic and non-Hispanic Black patients with comparable presentations. By contrast, disparities were attenuated or absent among patients deemed high risk by both nurses and the algorithm. Finally, analysis of the projected length-of-stay impact of substituting nurse-assigned with algorithmic triage scores suggests that algorithmic ESI decision support could reduce triage disparities with minimal effects on patient flow.

## 1 Main

Nearly two decades after the Institute of Medicine declared emergency departments (EDs) to be “at the breaking point” [1], overcrowding continues to strain U.S. EDs [2–4]. There were 155.4 million visits to U.S. EDs in 2022, roughly 47 per 100 people [5]. All ED visits begin with nurse triage, a process that assigns urgency and shapes downstream care (e.g. admission, length of stay). Consequently, even small, unwarranted variation in triage assignment may contribute to disparities in timeliness of care, length of stay, and mortality risk [6].

To reduce such variation, healthcare systems have adopted standardized triage protocols intended to support objective, repeatable assignments. The most widely used is the Emergency Severity Index (ESI) [7, 8], developed in 1998 and used in 94% of U.S. EDs as of 2019 [8]. ESI is a five-level scale intended to reflect urgency and risk of deterioration (1-immediate, 2-emergent, 3-urgent, 4-semi-urgent, 5-non-urgent). ESI is determined from clinical characteristics, including “high-risk” symptoms and “danger-zone” vital signs, and does not explicitly incorporate sex or race/ethnicity.

ESI is intended to standardize triage assessments, yet studies show frequent misapplication (59% accuracy [9]) and high inter-rater variability [10, 11]. Moreover, the process of assigning ESI scores may be susceptible to bias. Retrospective studies report that Black and Hispanic patients may be assigned less urgent scores than White patients [12–15], potentially contributing to longer waits, less comprehensive evaluation, misallocation of resources, and fewer admissions. Assignment practices also vary across sites, and the questions used to elicit and document patient status vary across triage nurses [16]. To reduce unwarranted variation, it is critical to identify where bias may enter the triage decision pathway, particularly at the boundary separating *high-acuity* patients (ESI 1–2) from *lower-acuity* patients (ESI 3–5), who can generally wait to be seen.

This requires identifying the conditions under which disparities in triage assignment arise. We hypothesized that ethnoracial disparities would be enriched in pathways that (1) carry less apparent clinical risk and (2) rely more heavily on subjective clinical judgment. We tested these hypotheses by implementing an automated ESI algorithm and generating algorithmic scores for approximately 1.4M encounters from four U.S. EDs over a 15-year period (Fig. 1). This enabled stratification of nurse-assigned triage by agreement with handbook criteria and finer separation of apparent high-versus low-risk visits. We then used causal inference methods to estimate the association between ethnoracial identity and the odds of receiving an emergency (ESI 2) nurse triage assignment, stratified by algorithmic ESI assessment.

**Fig. 1.**
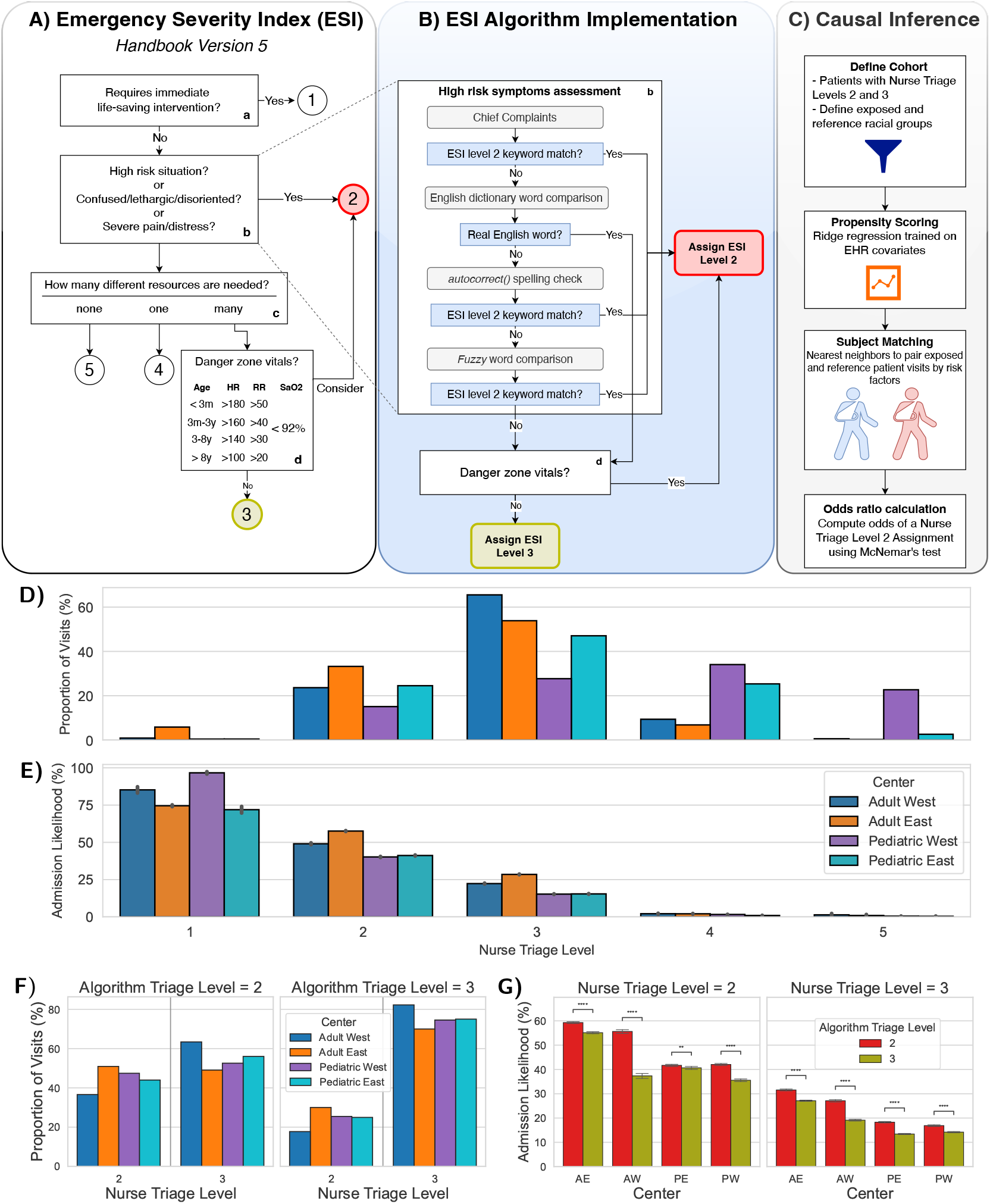
Emergency Severity Index (ESI) algorithm implementation and validation. **A)** Original ESI algorithm flowchart with triage levels 2 and 3 highlighted in red and green, respectively. **B)** Computational implementation of ESI algorithm levels 2 and 3, including high-risk symptom detection (box b) and danger-zone vital sign detection (box d). Algorithm ESI levels 2 and 3 are highlighted in red and green. **C)** Propensity score matching pipeline workflow. **D)** Distribution of nurse-assigned triage levels. Levels 2 and 3 are most common overall. **E)** Nurse-assigned triage level is associated with admission. Patients assigned high acuity (levels 1–2) are more likely to be admitted than those assigned levels 3–5. **F)** Concordance between algorithmic and nurse-assigned triage at levels 2 and 3. Roughly half of algorithmic level-2 encounters are assigned nurse level 3, whereas algorithmic level-3 encounters more closely agree with nurse assignments. **G)** Algorithmic ESI provides diagnostic information beyond nurse-assigned triage. Encounters assigned a more urgent algorithmic score than the nurse-assigned score are more likely to be admitted (right panel), and vice versa (left panel), across sites. *p*-value annotation legend: *: 1.00e-02 *< p <*= 5.00e-02; **: 1.00e-03 *< p <*= 1.00e-02; ***: 1.00e-04 *< p <*= 1.00e-03; ****: p *<*= 1.00e-04.

We analyzed 1,381,873 encounters at four EDs serving adults on the East (AE: 398,661 encounters) and West coasts (AW: 116,063 encounters) and children on the East (PE: 339,400 encounters) and West coasts (PW: 527,749 encounters). The cohort was diverse (30.8% Non-Hispanic White, 12.6% Non-Hispanic Black, 36.7% Hispanic, 5.1% Asian, and 10.4% Other); see Table S1. We focused on encounters assigned ESI levels 2 and 3, which comprise the majority of visits and represent the critical boundary between emergent (ESI 2) and urgent (ESI 3) care (Fig. 1E). This boundary is also strongly associated with admission likelihood (Fig. 1E) and is therefore a setting in which clinician judgment can meaningfully shape downstream care. Accordingly, the final cohort included encounters assigned ESI level 2 (AE: 33.2%, AW: 23.6%, PE: 24.5%, PW: 15.3%) or level 3 (AE: 53.9%, AW: 65.5%, PE: 47.1%, PW: 27.7%) at triage, totalling 342,232 (AE), 103,029 (AW), 242,734 (PE), and 226,864 (PW) encounters. Additional cohort details are provided in Table S1.

Comparison of nurse-assigned and algorithmic ESI scores showed substantial discordance (Fig. 1F), particularly for algorithmic ESI level 2, consistent with subjectivity at the level-2/3 boundary. To assess the validity of algorithmic assignments, we compared admission rates conditional on nurse-assigned and algorithmic ESI scores (Fig. 1G). Encounters assigned nurse level 2 were *less* likely to be admitted when the algorithm assigned level 3, and conversely, encounters assigned nurse level 3 were *more* likely to be admitted when the algorithm assigned level 2 (*p <* 6.1e-3, Mann–Whitney–Wilcoxon test, two-sided). These results support that the ESI handbook algorithm captures clinically meaningful severity information beyond nurse-assigned triage.

We estimated the odds of receiving a nurse-assigned ESI level 2, relative to propensity-matched Non-Hispanic White encounters, stratified by ethnoracial group and algorithmic triage level (Fig. 2). Across centers, Hispanic and Non-Hispanic Black patients generally had lower odds of nurse-assigned level 2 than matched Non-Hispanic White patients. For Hispanic patients, odds were lower at all centers (OR, AW: 0.91 [0.88–0.95]; AE: 0.75 [0.72–0.78]; PW: 0.85 [0.84–0.87]; PE: 0.83 [0.81–0.85]). Similarly, Non-Hispanic Black patients had lower odds at three of four centers (AW: 0.87 [0.80–0.95]; AE: 0.73 [0.71–0.74]; PE: 0.85 [0.81–0.85]).

**Fig. 2.**
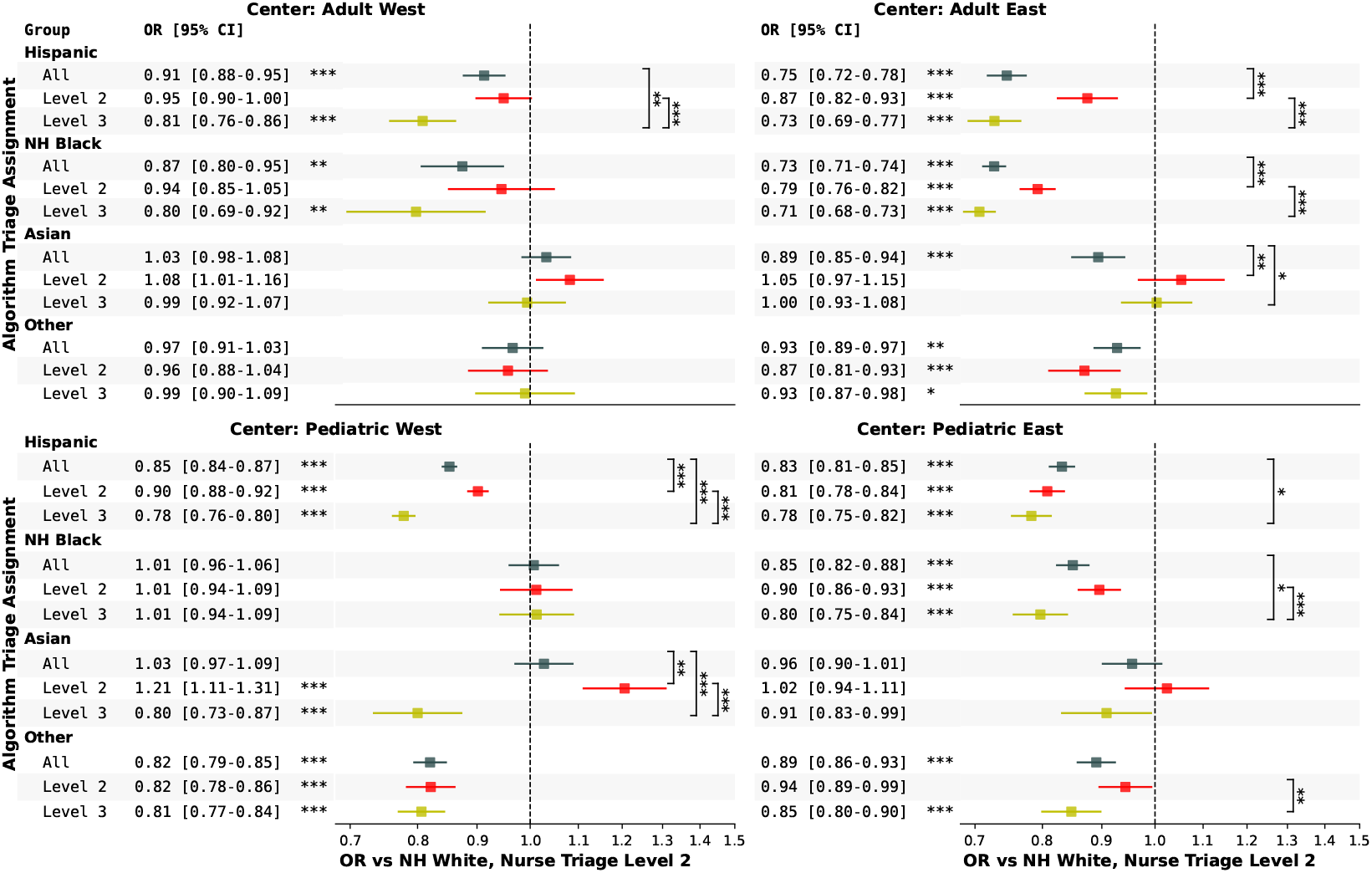
Ethnoracial disparities in emergency triage assignment by algorithmic ESI assessment. Odds ratios of receiving a nurse-assigned ESI level 2 (emergent) relative to Non-Hispanic White patients. Each subplot represents a care center (AW, AE, PW, PE). Results are shown for Hispanic, Non-Hispanic Black, Asian, and Other groups (y-axis). Results are further stratified by algorithmic triage assignment (y-axis). Adjusted odds ratios (ORs) are shown with 95% confidence intervals (CIs). Statistical significance of differences relative to Non-Hispanic White patients is denoted by asterisks in the test column accompanying the ORs. Additional within-group tests compare ORs across algorithmic triage levels and are denoted by vertical asterisks. *p*-value annotation legend: *: 1.00e-02 *< p <*= 5.00e-02; **: 1.00e-03 *< p <*= 1.00e-02; ***: 1.00e-04 *< p <*= 1.00e-03; ****: p *<*= 1.00e-04.

When stratified by algorithmic triage level, disparities were more pronounced among encounters assessed as lower risk (algorithmic level 3). Hispanic patients had reduced odds of high-acuity triage compared to matched Non-Hispanic White patients at all centers (AW: 0.81 [0.76–0.86]; AE: 0.73 [0.69– 0.77]; PW: 0.78 [0.76–0.80]; PE: 0.78 [0.75–0.82]). Non-Hispanic Black patients showed similar patterns at three of four centers (AW: 0.80 [0.69–0.92]; AE: 0.71 [0.68–0.73]; PE: 0.80 [0.75–0.84]). By contrast, among algorithmic level-2 encounters, disparities persisted but were generally attenuated for Hispanic patients at three centers (AE: 0.87 [0.82–0.93]; PW: 0.90 [0.88–0.92]; PE: 0.81 [0.78–0.84]). Non-Hispanic Black patients likewise showed reduced but persistent disparities at two centers (AE: 0.79 [0.76–0.82]; PE: 0.90 [0.86–0.93]). Overall, patients assessed as lower risk by the algorithm (level 3) exhibited larger disparities than those assessed as higher risk (level 2) across multiple centers (Hispanic: AW, AE, and PW *p <* 0.001; Non-Hispanic Black: AE and PE *p <* 0.001).

A more granular analysis of alignment between algorithmic ESI and nurse-assigned triage revealed site-specific differences in how ESI level-2 criteria were applied. The ESI algorithm first evaluates high-risk symptoms and then danger-zone vital signs when determining level 2. However, centers differed in the relative contribution of these factors to nurse-assigned level-2 decisions (Fig. S1). In the Western adult and pediatric centers (AW and PW), nurses more often assigned level 2 in the presence of danger-zone vital signs than high-risk symptoms. Conversely, in the Eastern centers (AE and PE), nurses more frequently assigned level 2 based on high-risk symptoms than danger-zone vital signs.

A key translational question is whether algorithmic ESI scores could serve as clinical decision support to reduce disparities while preserving patient flow. To probe this, we estimated the impact on ED length of stay (LOS) if algorithmic ESI scores were used in place of nurse-assigned triage scores. We used an accelerated failure time model to estimate the expected change in ED LOS under replacement of nurse-assigned with algorithmic scores, evaluated at triage. At sites AW, AE, PW, and PE, algorithmic ESI would change the proportion of level-2 assignments by 75.7%, -0.1%, 30.5%, and 44.3%, and change the proportion of level-3 assignments by -27.3%, 0.1%, -16.9%, and -23.1%, respectively. Replacing nurse-assigned scores with algorithmic scores was estimated to increase mean ED LOS by 2.0% [2.0–2.1], 1.0% [0.9–1.0], 3.4% [3.4–3.5], and 4.6% [4.5–4.7] at sites AW, AE, PW, and PE, respectively. In AW, where time to admit/discharge decision is available, the estimated change in time to disposition was negligible (-0.1% [-0.1–0.0]). Given the high prevalence of level-3 encounters, a large relative increase in level-2 assignments can be partially offset by a small relative decrease in level-3 assignments. In practice, an ESI algorithm would be decision support rather than a replacement for clinician judgment, and these results suggest potential to reduce disparities with modest or negligible impact on ED throughput.

In summary, we observed significant ethnoracial disparities in triage assignment across four diverse emergency departments, with under-prioritization of minoritized patients relative to Non-Hispanic White patients across centers. These disparities persisted after adjustment for clinical, demographic, and hospital factors available at triage. Notably, disparities were most pronounced among encounters assessed as lower urgency by the ESI algorithm. These findings indicate that disparities are concentrated in discretionary triage decisions where clinicians may be more willing to (differentially) extend the benefit of the doubt, particularly for patients assessed as lower risk by standardized criteria.

Beyond documenting disparities, these findings have important implications for emergency care practice and quality improvement. Our results suggest that the ESI framework, while standardized in principle, is operationalized in ways that allow subjective judgment to differentially influence triage decisions at key decision boundaries. Algorithmic or rules-based decision support that explicitly encodes ESI criteria could therefore serve as a tool to promote consistency in triage assignment, particularly for patients whose presentations fall near acuity thresholds.

Such tools could be integrated into triage workflows as decision aids rather than replacements for clinical judgment, and may inform updates to triage training, audit processes, and quality assurance programs by identifying systematic deviations from guideline-based criteria. More broadly, our findings highlight the need for triage systems and clinical practice guidelines to be evaluated not only for overall accuracy and efficiency, but also for their equity impacts in real-world implementation. Careful design, monitoring, and governance of decision support tools will be essential to ensure that efforts to standardize triage reduce—rather than entrench—existing disparities at the point of first contact in emergency departments.

## 2 Methods

Here we describe ESI algorithm development, the causal inference approach to estimating disparities in triage assignment, and data preparation for the study.

### 2.1 ESI Algorithm Development

#### 2.1.1 High-risk patient identification

According to the ESI handbook [8], level-2 criteria include conditions that may rapidly deteriorate or require time-sensitive treatment (Fig. 1A). To computationally replicate this, we extracted 104 high-risk keywords from the ESI handbook and matched them to ED chief complaints (Fig. 1B).

#### 2.1.2 Danger zone vitals identification

When high-risk symptoms are absent but multiple resources are anticipated, ESI next considers vital signs (Fig. 1A). This assessment includes heart rate, respiratory rate, and oxygen saturation. If any vital sign exceeds the danger-zone threshold and multiple resources are anticipated, ESI recommends escalation to level 2 [8].

To implement this component, we stratified patients into age groups and compared triage vital signs with age-specific danger-zone thresholds. If any of the three vital signs exceeded threshold, the algorithm assigned level 2. Recent versions of the ESI handbook introduce a “consider” factor [8], allowing clinician discretion when danger-zone vital signs are present, whereas earlier versions did not [7]. We followed the original ESI definition, in which any danger-zone vital sign results in automatic assignment to level 2. We further examine this in Fig. S1.

### 2.2 Statistical Analysis

We used causal inference methods to estimate the association between patient race/ethnicity and the odds of receiving an emergency (ESI 2) nurse triage assignment. We approximated the causal effect by matching encounters on clinical presentation and contextual factors before estimating adjusted odds ratios. The approach is described below.

#### 2.2.1 Exposure of interest and outcome measure

The primary exposure was patient race/ethnicity, categorized as Hispanic, Non-Hispanic Black, Asian, and Other. Encounters with race/ethnicity recorded as “Unknown” were excluded (3.8% on average across datasets) due to uncertainty and potential misclassification. The primary outcome was assignment to nurse-assigned ESI level 2 at triage.

#### 2.2.2 Adjustment variables

We adjusted for variables available at triage and, where available, diagnoses recorded during the visit to support clinically meaningful matching. All datasets included sex, age, arrival mode, number of prior visits (with and without admission), comorbidity index, triage vital signs (heart rate, respiratory rate, oxygen saturation), and chief complaint. PE and PW additionally included preferred language, social deprivation index, miles traveled, state of origin, number of patients at arrival, and weight. Temporal variables (year and time of arrival) were available for PE and PW, and insurance information was available for PE, PW, and AW.

### 2.3 Odds ratio calculation

To assess disparities in ESI level assignment, we performed propensity score matching and estimated adjusted odds ratios [17], using Non-Hispanic White patients as the reference group because it was the largest group overall. For each comparison, analyses were restricted to encounters from the focal ethnoracial group and the reference group. Propensity scores were estimated using logistic regression with ridge regularization, incorporating covariates described in Section 2.2.2. Matching was performed using a ball-tree nearest-neighbors algorithm [18], with calipers of 10% and 20% of the standard deviation of fitted propensity scores. This approach achieved covariate balance across groups, with absolute standardized mean differences ≤ 0.1 for all covariates [17].

Odds ratios were computed in matched samples using McNemar’s test with Bonferroni correction for multiple comparisons following 1:1 nearest-neighbor matching. This test assumes independence between matched pairs and conditional independence of outcomes within pairs given the matching variables, assumptions that are reasonable given the encounter-level analysis and strict matching criteria. Differences across algorithmic triage strata within an ethnoracial group were assessed using a z-test, which assumes approximate normality of the log-odds ratio estimates and independent variance estimates across strata. Given the large sample sizes within strata, these normal approximation and variance assumptions are expected to hold.

### 2.4 Dataset Preparation

Each dataset was filtered for quality control (see Table S2). Visit records were discarded if any of the following criteria were met: unknown sex (i.e. sex recorded as U or X); missing demographic information; missing patient identifier; disposition other than admission or discharge (e.g. death, left without being seen, left against medical advice), or missing disposition with no associated vitals (suggesting departure before triage); missing chief complaint or primary visit diagnosis for datasets where these were recorded separately (suggesting data loss rather than inability to report a complaint); implausibly long visit length in the context of the dataset (generally ≥ two weeks), suggesting a recording error; irreconcilable timestamps (e.g. departure time earlier than arrival time).

Variables were processed into categories as described in Table S3. We aimed for consistency across datasets but were constrained by site-specific recording practices. For example, PE recorded both sex and gender, PW and AE recorded only sex, and AW recorded only gender, which was not defined and was assumed to reflect provider-recorded sex. Race/ethnicity recording also differed; notably, AE defined Hispanic’, White’, and ‘Black’ as mutually exclusive categories, whereas other sites recorded Hispanic ethnicity separately from race. We harmonized categories as Hispanic, Non-Hispanic Black, Non-Hispanic Asian (denoted Asian”), Non-Hispanic White, Non-Hispanic Other (denoted Other”), and “Unknown”. Temporal variables (e.g. month, year, crowding) were unavailable for adult datasets, which obscured visit dates for deidentification. In AW, age was shifted by up to two years (all patients were ≥ 18 years, so this shift does not affect ESI algorithm assignments). Both adult datasets truncated ages above 90 at first visit; because such patients were rare, we coarsened older ages into an ‘80+’ group.

Where available, socio-economic and geographic measures (e.g. miles traveled) were divided into dataset-specific quartiles. Socio-economic deprivation scores were obtained from the Robert Graham Center database (2019; most recent available) [19]. This resource assigns a score from 1 (least deprived) to 100 (most deprived) to each U.S. zip code. *Distance traveled to the hospital* was computed assuming travel from the patient’s home address.

Chief complaints were grouped using adult and pediatric schemas from [20] and [21], respectively, with site-specific additions where needed to capture common reasons for visit (e.g. shingles for AW; see preprocessing code for details). Primary diagnoses were transformed into binary indicators corresponding to Charlson Comorbidity Index categories for adult datasets (using the R package comorbidity) and the Pediatric Comorbidity Index [22] for pediatric datasets. Where available, prior diagnoses (comorbidities) were encoded similarly (e.g. prior diabetes, congestive heart failure, renal disease, cancer). For AE, where prior diagnoses were unavailable, we extracted therapeutic medication classes at the time of visit as proxies for chronic conditions, yielding 85 binary indicators (e.g. thyroid therapy).

## Data Availability

The Adult West cohort data is available online at https://physionet.org/content/mc-med/1.0.0/. The Adult East cohort is available online at https://physionet.org/content/mimic-iv-ed/2.2/. The Pediatric East and West cohorts are not available under the terms of the institutional IRBs. Certain data produced in the present study may be made available upon reasonable request to the authors. Code supporting this analysis is available from https://github.com/cavalab/ESI. The repository includes
scripts for preparing the datasets for analysis, as well as the analysis scripts themselves.

## Supplementary information

Code supporting this analysis is available at https://github.com/cavalab/ESI. The repository includes scripts for dataset preparation and analysis. Supplementary materials include additional cohort descriptions, analyses of the ESI algorithm, and tables documenting preprocessing decisions.

## Acknowledgements

Research reported in this publication was supported by the National Library of Medicine of the National Institutes of Health (NIH) Award R01LM014300. The content is the responsibility of the authors and does not necessarily represent the official views of NIH.

