## Supplement for "The Benefit of the Doubt Phenomenon in Emergency Triage Assignment Disparities"

### S1 Code Availability

Code supporting this analysis is available from https://github.com/cavalab/ESI. The repository includes scripts for preparing the datasets for analysis, as well as the analysis scripts themselves.

### S2 Supplemental Results

#### S2.1 Cohort Description

Table S1 provides detailed cohort information for the four medical center EDs we analyzed in this study.

#### S2.2 Extended analysis of triage algorithm

The ESI algorithm contains two paths that arrive at triage level 2 (see Fig. 1). The first is through the presentation of high risk symptoms. The second is through the presence of concerning (i.e., "danger zone") vitals, in which case the handbook recommends reconsidering a triage level 2 assignment, even absent high risk symptoms.

Both routes can be seen as relying on some degree of clinical judgement. High risk symptoms are interpreted by the clinican from the patient or their guardian, which creates the potential for subjective translation. Meanwhile, the ESI version 5 handbook explicitly instructs clinicians to use their judgement when considering whether to classify patients as high risk given their vitals (earlier ESI versions always recommend assigning triage 2). Thus, an open question we sought to answer was which route was more often used to assign triage level 2, and whether the routes exhibited markedly different disparities in triage assignment.

We found that there was a significant West coast/East coast divide in which paths led to nurse triage level 2 assignments (see Fig. S1). The adult and pediatric west coast sites were significantly more likely to assign triage 2 when patients had danger zone vitals (median [IQR] LR+, AW: 1.72 [1.62-1.79]; PW: 1.49 [1.48 - 1.52]), versus when patients had high risk symptoms (LR+, AW: 0.98 [0.93-1.06]; PW: 1.34 [1.14 - 1.42]). Conversely, adult and pediatric east coast sites were much more likely to assign triage 2 for patients with high risk symptoms (LR+, AE: 1.68 [1.63 - 1.83]; PE: 1.34 [1.14 - 1.42]); on Eastern sites, danger zone vitals had little impact on the likelihood of assignment themselves (LR+, AE: 1.11 [1.08 - 1.13]; PE: 1.09 [0.98 - 1.17]).

In all sites, patients with *both* high risk symptoms and danger zone vitals had the highest likelihood ratio of triage 2 assignments compared to patients with one or the other (median LR+: 1.76 - 2.16). This result suggests, in all sites, that the ESI algorithm is being used in a different manner than it was designed. By following the algorithm, *a priori*, we would expect danger zone vitals to only impact the likelihood of triage 2 assignments when patients do not express high risk symptoms. Furthermore, we would expect high risk symptoms, regardless of danger zone vitals, to independently increase the likelihood of triage 2 assignment. By contrast, in AW, high risk symptoms alone do not increase the likelihood of triage 2 assignments; similarly, in the East coast sites, danger zone vitals alone do not increase triage 2 assignment likelihood ratios. Yet danger zone vitals paired with high risk symptoms increase the likelihood of triage 2 assignment above and beyond either observation alone, suggesting patient reported symptoms and vitals are often considered in concert, rather than serially, when making triage decisions. In other words, all sites appear to consider both hallmarks of triage 2 assignment, but a) consider them differently depending on the site and 2) depart from the guidelines by the manner in which they combine these risk factors.

There were some small differences in likelihood ratios by patient race but in general they were consistent. These findings highlight that site-specific differences in triage are significant, and complicate efforts to apply a one-size-fits-all solution to the problem of triage assignment disparities.

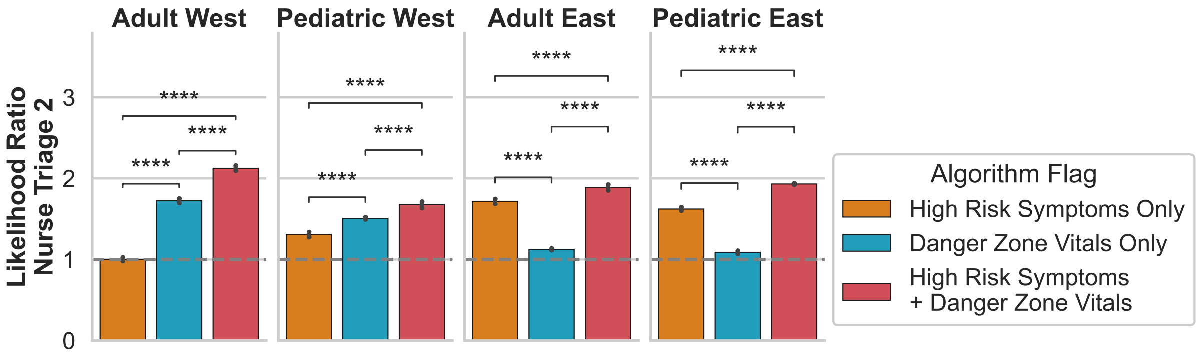


**Fig. S1 Variation in factors driving Nurse Triage Assignment among Centers.** The ESI triage algorithm relies first on high risk symptoms, then danger zone vitals, to assign triage level 2. The Western and Eastern centers differ in the likelihood of a Nurse Triage 2 assignment based on these factors. In the adult and pediatric western centers, Nurse Triage Level 2 assignments are more likely when patients have danger zone vitals than when they have high risk symptoms; in the Eastern centers, we observe the opposite. Asterisks denote the result of paired Mann-Whitney-Wilcoxon two-sided tests. $p$-value annotation legend: ns: 5.00e-02 $< p <=$ 1.00e+00; *: 1.00e-02 $< p <=$ 5.00e-02; **: 1.00e-03 $< p <=$ 1.00e-02; ***: 1.00e-04 $< p <=$ 1.00e-03; ****: p <= 1.00e-04.

**Acknowledgements.** Research reported in this publication was supported by the National Library of Medicine of the National Institutes of Health (NIH) Award R01LM014300. The content is the responsibility of the authors and does not necessarily represent the official views of NIH.

**Table S1** Descriptive statistics for the Emergency Departments used in the study.

| | | | Grouped by Center | | | |
|---|---|---|---|---|---|---|
| | | Overall | Adult East | Adult West | Pediatric East | Pediatric West |
| Visits, n | | 1,381,873 | 398,661 | 116,063 | 339,400 | 527,749 |
| Unique Patients, n | | 685,493 | 196,421 | 69,402 | 179,944 | 239,726 |
| Age Group, n (%) | < 3 mo. | 35,085 (2.5) | | | 14,178 (4.2) | 20,907 (4.0) |
| | 3-6 mo. | 21,869 (1.6) | | | 9,332 (2.7) | 12,537 (2.4) |
| | 6-12 mo. | 56,326 (4.1) | | | 20,089 (5.9) | 36,237 (6.9) |
| | 12-18 mo. | 52,066 (3.8) | | | 18,300 (5.4) | 33,766 (6.4) |
| | 1.5-3 | 115,141 (8.3) | | | 41,084 (12.1) | 74,057 (14.0) |
| | 3-5 | 115,520 (8.4) | | | 40,078 (11.8) | 75,442 (14.3) |
| | 5-10 | 197,352 (14.3) | | | 69,520 (20.5) | 127,832 (24.2) |
| | 10-15 | 142,313 (10.3) | | | 58,468 (17.2) | 83,845 (15.9) |
| | 15+ | 131,477 (9.5) | | | 68,351 (20.1) | 63,126 (12.0) |
| | 18-29 | 88,749 (6.4) | 70,525 (17.7) | 18,224 (15.7) | | |
| | 30-39 | 68,095 (4.9) | 50,214 (12.6) | 17,881 (15.4) | | |
| | 60-69 | 81,313 (5.9) | 63,552 (15.9) | 17,761 (15.3) | | |
| | 70-79 | 63,636 (4.6) | 48,618 (12.2) | 15,018 (12.9) | | |
| | 40-49 | 64,769 (4.7) | 49,346 (12.4) | 15,423 (13.3) | | |
| | 50-59 | 84,787 (6.1) | 67,493 (16.9) | 17,294 (14.9) | | |
| | 80+ | 63,375 (4.6) | 48,913 (12.3) | 14,462 (12.5) | | |
| Sex, n (%) | F | 687,275 (49.7) | 216,552 (54.3) | 63,042 (54.3) | 161,725 (47.7) | 245,956 (46.6) |
| | M | 694,598 (50.3) | 182,109 (45.7) | 53,021 (45.7) | 177,675 (52.3) | 281,793 (53.4) |
| Race, n (%) | Asian | 70,288 (5.1) | 17,661 (4.4) | 18,945 (16.3) | 13,260 (3.9) | 20,422 (3.9) |
| | Hispanic | 506,986 (36.7) | 32,458 (8.1) | 31,943 (27.5) | 88,834 (26.2) | 353,751 (67.0) |
| | NH Black | 174,044 (12.6) | 86,921 (21.8) | 7,296 (6.3) | 50,853 (15.0) | 28,974 (5.5) |
| | NH White | 425,064 (30.8) | 232,435 (58.3) | 43,101 (37.1) | 116,339 (34.3) | 33,189 (6.3) |
| | Other | 144,290 (10.4) | 23,622 (5.9) | 13,839 (11.9) | 33,639 (9.9) | 73,190 (13.9) |
| | Unknown | 61,201 (4.4) | 5,564 (1.4) | 939 (0.8) | 36,475 (10.7) | 18,223 (3.5) |
| Triage Acuity, n (%) | 1.0 | 27,887 (2.0) | 22,975 (5.8) | 1,029 (0.9) | 1,535 (0.5) | 2,348 (0.4) |
| | 2.0 | 321,814 (23.4) | 130,564 (33.2) | 27,312 (23.6) | 83,163 (24.5) | 80,775 (15.3) |
| | 3.0 | 593,045 (43.1) | 211,668 (53.9) | 75,717 (65.5) | 159,571 (47.1) | 146,089 (27.7) |
| | 4.0 | 302,683 (22.0) | 26,855 (6.8) | 10,830 (9.4) | 85,969 (25.4) | 179,029 (33.9) |
| | 5.0 | 129,673 (9.4) | 955 (0.2) | 668 (0.6) | 8,811 (2.6) | 119,239 (22.6) |
| Disposition, n (%) | Admitted | 309,794 (22.4) | 157,514 (39.5) | 31,574 (27.2) | 60,843 (17.9) | 59,863 (11.3) |
| | Discharged | 1,069,915 (77.4) | 241,147 (60.5) | 84,489 (72.8) | 278,557 (82.1) | 465,722 (88.2) |
| | Transfer | 2,164 (0.2) | | | | 2,164 (0.4) |

| Site | Number of unique visits | | Number of unique patients | |
|---|---|---|---|---|
| | Before filtering | After filtering (% remaining) | Before filtering | After (% remaining) |
| Adult West | 118,385 | 116,063 (98.0%) | 70,545 | 69,402 (98.4%) |
| Adult East | 425,087 | 398,661 (93.8%) | 205,504 | 196,421 (95.6%) |
| Pediatric West | 582,169 | 527,749 (90.7%) | 255,907 | 239,726 (93.7%) |
| Pediatric East | 352,324 | 339,400 (96.3%) | 184,086 | 179,944 (97.7%) |

**Table S2** A description of the size of each dataset before and after the filtering process.

**Table S3** A description of the categorisation of variables in each of the four datasets. H: Hispanic, NH: Non-Hispanic, W: White, B: Black; CCI: Charlson comorbidity index; PCI: pediatric comorbidity index; st. dev.: standard deviation. Q1, Q4, etc. refer to dataset-specific quartiles.

| Variable | Adult West | Adult East | Pediatric West | Pediatric East |
|---|---|---|---|---|
| Sex | M, F | M, F | M, F | M, F (trans or NB gender identity separately recorded) |
| Race | Asian, HB, HW, H, NHB, NHW, Other, Unknown | Asian, H, NHB, NHW, Other, Unknown | Asian, H, HW, NHB, NHW, Other, Unknown | Asian, H, HW, NHB, NHW, Other, Unknown |
| Age | 18-29, 30-39, ... 70-79, 80+ | Same as AW | 0-3m, 3-6m, 6-12m, 12-18m, 18-36m, 3-5y, 5-10y, 10-15y, 15y+ | Same Same as PW |
| Insurance | Medicaid, Medicare, Other/Unknown | N/A | Public, Private, Unknown | Public, Private |
| Triage acuity (ESI score) | 1, 2, 3, 4, 5, unknown | | | |
| Mode of arrival | EMS, Self, Other/Unknown | | | |
| Visit history | Number of admissions/discharges from the ED in the last 30 days | | | |
| Primary language | N/A | N/A | Armenian, English, Mandarin, Russian, Spanish, Other/Unknown | Arabic, Cape Verdean, English, Haitian Creole, Mandarin, Portuguese, Spanish, Other |
| Socio-economic deprivation | N/A | N/A | Q1, Q2, Q3, Q4, unknown | Q1, Q2, Q3, Q4, unknown |
| Miles travelled | N/A | N/A | Q1, Q2, Q3, Q4, unknown | Q1, Q2, Q3, Q4, unknown |
| State of origin | N/A | N/A | In-state, out-of-state, unknown | In-state, out-of-state, unknown |
| Chief complaint | Categorized per [1] | Categorized per [1] | Categorized per [2] | Categorized per [2] |
| Comorbidities | Based on CCI | Taken from common medications | Based on PCI | Based on PCI |
| Visit diagnosis | Based on CCI | Based on CCI | Based on PCI | Based on PCI |
| Weight | N/A | N/A | Number of st. devs. from age-group mean | Same as PW |
| Time of arrival | N/A | N/A | Weekend (yes/no), time of day at arrival (0-6h, 6-12h, ..), season (Dec-Feb, Mar-May...) | Same as PW |
| Crowding | N/A | N/A | Number of other patients waiting in the ED at the time of arrival | Same as PW |
